# PanCanAID – Pancreas Cancer Artificial Intelligence Driven Diagnosis in CT Scan Imaging: A Protocol for a Multicentric Ambispective Diagnostic Study

**DOI:** 10.1101/2023.08.03.23293596

**Authors:** Seyed Amir Ahmad Safavi-Naini, Armin Behnamnia, Faezeh Khorasanizadeh, Ali Soroush, Farhad Zamani, Faeze Salahshour, Amir Sadeghi, Seyedmahdi Mirtajaddini, Ashkan Zandi, Fatemeh Shojaeian, Maryam Saeedi, Azade Ehasni, Abdolhamid Chavoshi Khamneh, Zhaleh Mohsenifar, Farid Azmoudeh Ardalan, Kavous Firouznia, Shabnam Shahrokh, Masoomeh Raoufi, Pooneh Dehghan, Pardis Ketabi Moghadam, Alireza Mansour-Ghanaei, Parinaz Mellatdoust, Habib Malekpour, Alireza Rasekhi, Fariborz Mansour-Ghanaei, Masoudreza Sohrabi, Fariba Zarei, Amir Reza Radmard, Hossein Ghanaati, Hamid Assadzadeh Aghdaei, Mohammad Reza Zali, Hamid R. Rabiee

**Author notes:** Corresponding to (HRR) & (HAA) & (HG) & (FZ) & (MS) & (FMG).

## Abstract

**Introduction:** Pancreatic cancer is thought to have an extremely dismal prognosis. Most cancer-related deaths occur from metastasis rather than the primary tumor, although individuals with tumors smaller than 1 cm in diameter have more than 80% 5-year survival. Thus, the current protocol introduces PanCanAID project which intends to develop several computer-aided-diagnosis (CAD) systems to enhance pancreatic cancer diagnosis and management using CT scan imaging.

**Methods and analysis:** Patients with pathologically confirmed pancreatic ductal adenocarcinoma (PDAC) or pancreatic neuroendocrine tumor (PNET) will be included as pancreatic cancer cases. The controls will be patients without CT evidence of abdominal malignancy. A data bank of contrast-enhanced abdominopelvic CT scans, survival data, and demographics will be collected from ten medical centers in four provinces. Endosonography images and clinical data, if available, will be added to the data bank. Annotation and manual segmentation will be handled by radiologists and confirmed by a second expert radiologist in abdominal imaging. PanCanAID intelligent system is designed to (1) detect abdominopelvic CT scan phase, (2) segment pancreas organ, (3) diagnose pancreatic cancer and its subtype in arterial phase CT scan, (4) diagnose pancreatic cancer and its subtype in non-contrast CT scan, (5) carry out prognosis (TNM stage and survival) based on arterial phase CT scan, (6) and estimate tumor resectability. A domain adaptation step will be handled to use online data and provide pancreas organ segmentation to reduce the segmentation time. After data collection, a state-of-the-art deep learning algorithm will be developed for each task and benchmarked against rival models.

**Conclusion:** PanCanAID is a large-scale, multidisciplinary AI project to assist clinicians in diagnosing and managing pancreas cancer. Here, we present the PanCanAID protocol to assure the quality and replicability of our models. In our experience, the effort to prepare a detailed protocol facilitates a positive interdisciplinary culture and the preemptive identification of errors before they occur.

## Introduction

Among all cancer types, pancreatic cancer has an especially dismal prognosis (1). At the time of presentation, only 11% of patients are at an early enough stage to qualify for curative surgery (1–3). In particular, individuals with tumors smaller than 1 cm in diameter showed a relatively favorable average long-term survival rate of 80.4% at five years (4). Therefore, effective early detection of pancreatic cancer is critical for increasing the proportion of individuals who can qualify for treatments that reduce mortality (1). Current methods of pancreatic cancer detection include abdominal computed tomography (CT), endoscopic ultrasonography (EUS), endoscopic retrograde cholangiopancreatography (ERCP), and magnetic resonance imaging (MRI) (5).

**Fig 1.**
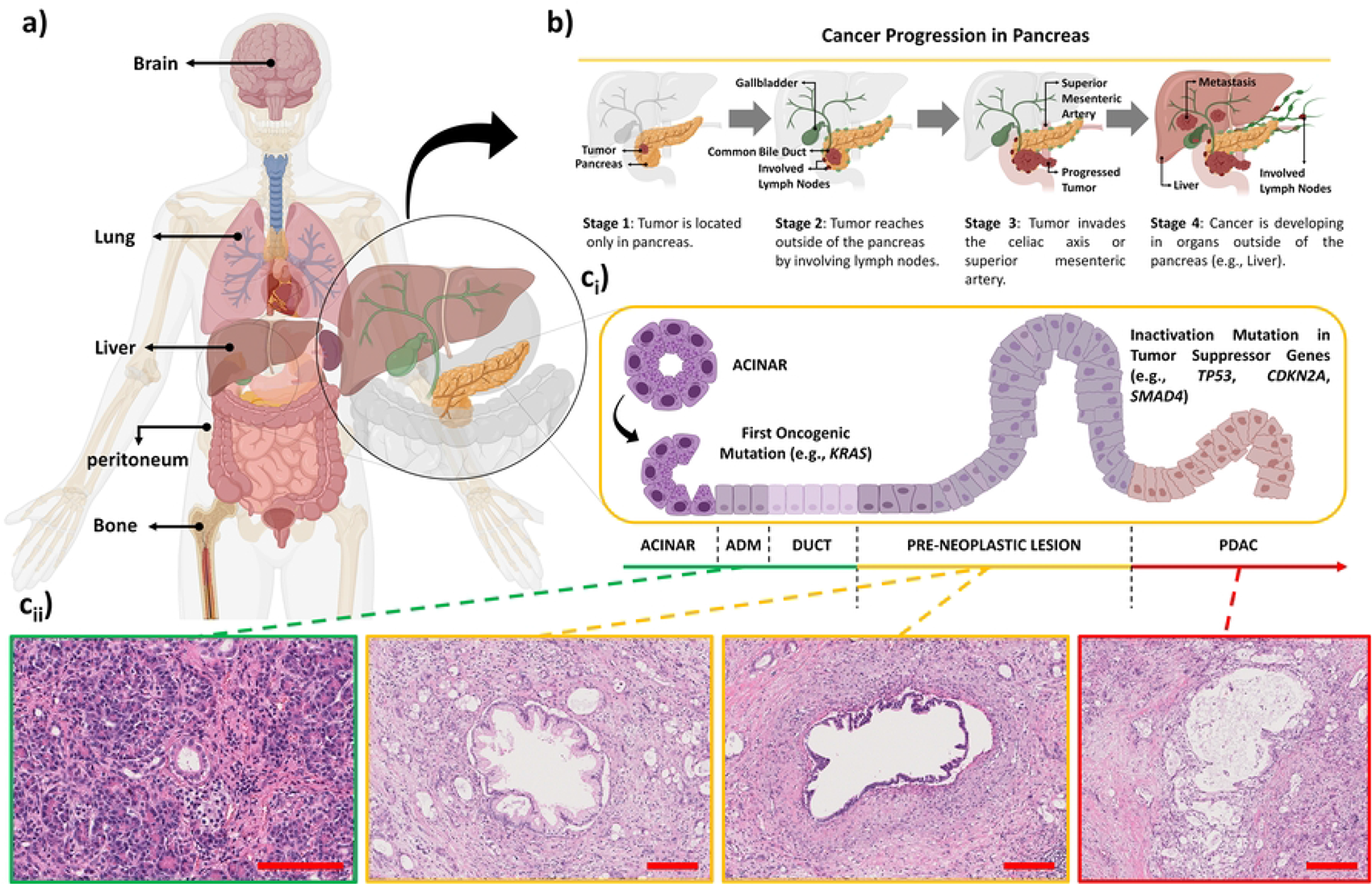
a) Pancreatic cancer can metastasize to several secondary sites, including the liver, lung, peritoneum, bone, and brain. The most common secondary site is the liver, which is affected in more than half of the cases of metastatic pancreatic cancer. The lung is the second most common site, followed by the peritoneum (6). b) Cancer progression in the pancreas. The tumor is initially confined to the pancreas in stage #1, but it spreads beyond the pancreas to involve nearby lymph nodes in stage #2. By stage #3, the tumor has invaded either the celiac axis or the mesenteric artery. In stage #4, cancer involves other organs outside the pancreas (7). ci) Pancreatic cancer often results from a sequence of genetic mutations that transform normal pancreatic mucosa into an invasive malignancy through precursor lesions. The three most widely recognized precursor lesions are Pancreatic Intraepithelial Neoplasia (PanIN), Intraductal Papillary Mucinous Neoplasm (IPMN), and Mucinous Cystic Neoplasm (MCN). PanIN is the most common precursor lesion for pancreatic ductal adenocarcinoma (PDAC), and genetic abnormalities found in these lesions are common in adjacent PDAC. The histological pattern of PanIN progression also reflects the accumulation of mutations in cancerous tissue. KRAS mutations and shortened telomeres characterize low-grade PanIN lesions. At the same time, high-grade PanIN and PDAC tissues display mutations in p16, p53, CDNK27, and SMAD4, along with a higher frequency of KRAS mutation (8). cii) Image with a green border shows normal acini and normal ducts. The base of acinar cells turns blue due to the abundance of RNA and nuclei, while the cells’ apex (or lumenal aspect) appears pink due to the high presence of zymogen proteins that function as digestive enzymes. Intralobular ductule in cross section is obvious. The ductule’s lumen contains a granular proteinaceous precipitate that appears pink due to pancreatic juice. The nuclei in images with orange borders are enlarged, hyperchromatic, and show moderate to severe nuclear atypia, with prominent nucleoli representing PanIN. The cytoplasm may be abundant and mucin filled. PanIN, a pre-neoplastic lesion of the pancreas, is classified into low-grade and high-grade based on the degree of dysplasia. The epithelial cells display severe nuclear atypia and anaplasia, with loss of polarity and increased mitotic activity. Image with a red border, PDAC is the most common form of pancreatic cancer. On H&E staining, PDAC lesions typically exhibit the following features: desmoplastic reaction, hyperchromatic nuclei with irregular contours and clumped chromatin tumor cells, mitotic figures, and invasion.

CT scans and EUS are the commonly used imaging examinations for pancreas cancer (9, 10). EUS offers an excellent spatial resolution of the pancreas, and CT scans give information about the tumor and its relationship to surrounding structures. However, EUS is an invasive procedure, and its performance depends on the endoscopist’s skill (11, 12). Radiologists also require a considerable amount of training to identify early-stage pancreatic tumors. In a retrospective analysis of pancreatic cancer cases, tumors were detectable in CT images three years before clinical diagnosis (13). Even with expert radiologists, exhaustion and negligence can additionally lead to missed diagnoses (14). These findings suggest an improved review of CT scan exams could increase the proportion of pancreatic cancers diagnosed early.

Contrast-enhanced CT scan (CECT) is the preferred technique for pancreas imaging since it characterizes the tumor and surrounding tissue. After the injection of intravenous contrast (IV), the operator takes sequential CT scans at 45 seconds (late arterial phase) and 60 seconds (portovenous phase). Some centers obtain more detailed triple-phase “pancreas protocol” imaging (arterial, venous, and portal) to improve visualization of the tumor and characterization of invasion (15). The effective interpretation of multiple collections of images in different phases requires significant experience and attention.

Optimism has steadily grown over the potential of computer-assisted radiology techniques to facilitate early diagnosis and timely management (16, 17). Machine learning (ML) models can explicitly explore hidden patterns in the data and have produced groundbreaking results in almost all fields of medical imaging (18). Expert radiologists often outperform ML models.

However, the cost and limited availability of such expertise impair the early detection of pancreatic cancer (18). ML-driven computerized clinical decision support systems (CDSS) can help less experienced clinicians decrease time-to-diagnosis, increase accuracy, reduce interobserver variability, promote equitable healthcare access, and enhance cost-effectiveness (14, 18).

A sizeable multicenter image dataset and interdisciplinary framework are required to develop a generalizable and practical CDSS. Both requirements are especially challenging for pancreatic cancer as the disease is rare, and its management is a winding journey involving many points of care (17, 19). The challenge of obtaining data could explain why few high-performance but data-demanding deep learning models have been published for pancreatic cancer (20). As of 2023, 2 out of 13 studies of ML-based pancreatic cancer detection on CT scan imaging had more than 300 participants (21), some of which have been summarized in **Table 1**. Although artificial intelligence (AI)-assisted pancreatic cancer detection is rapidly growing, studies have been hampered by small sample sizes and lack of external validation. Besides, new approaches such as Segment Anything Model (SAM) may improve model performance and facilitate data labeling (20–22).

**Table 1.**
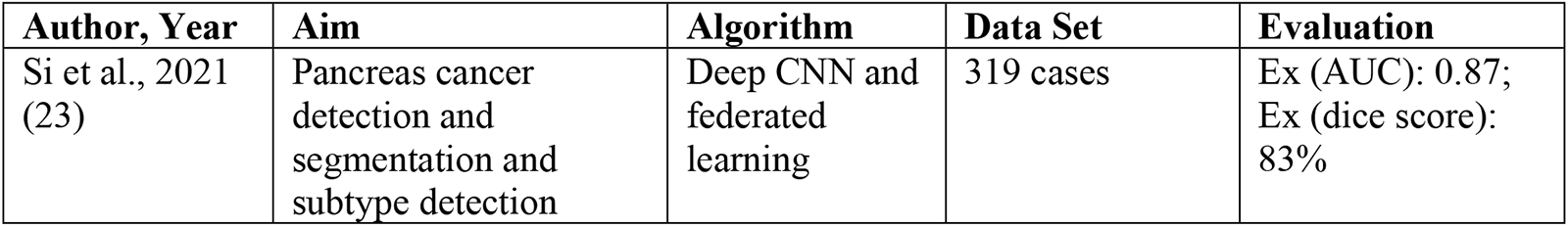
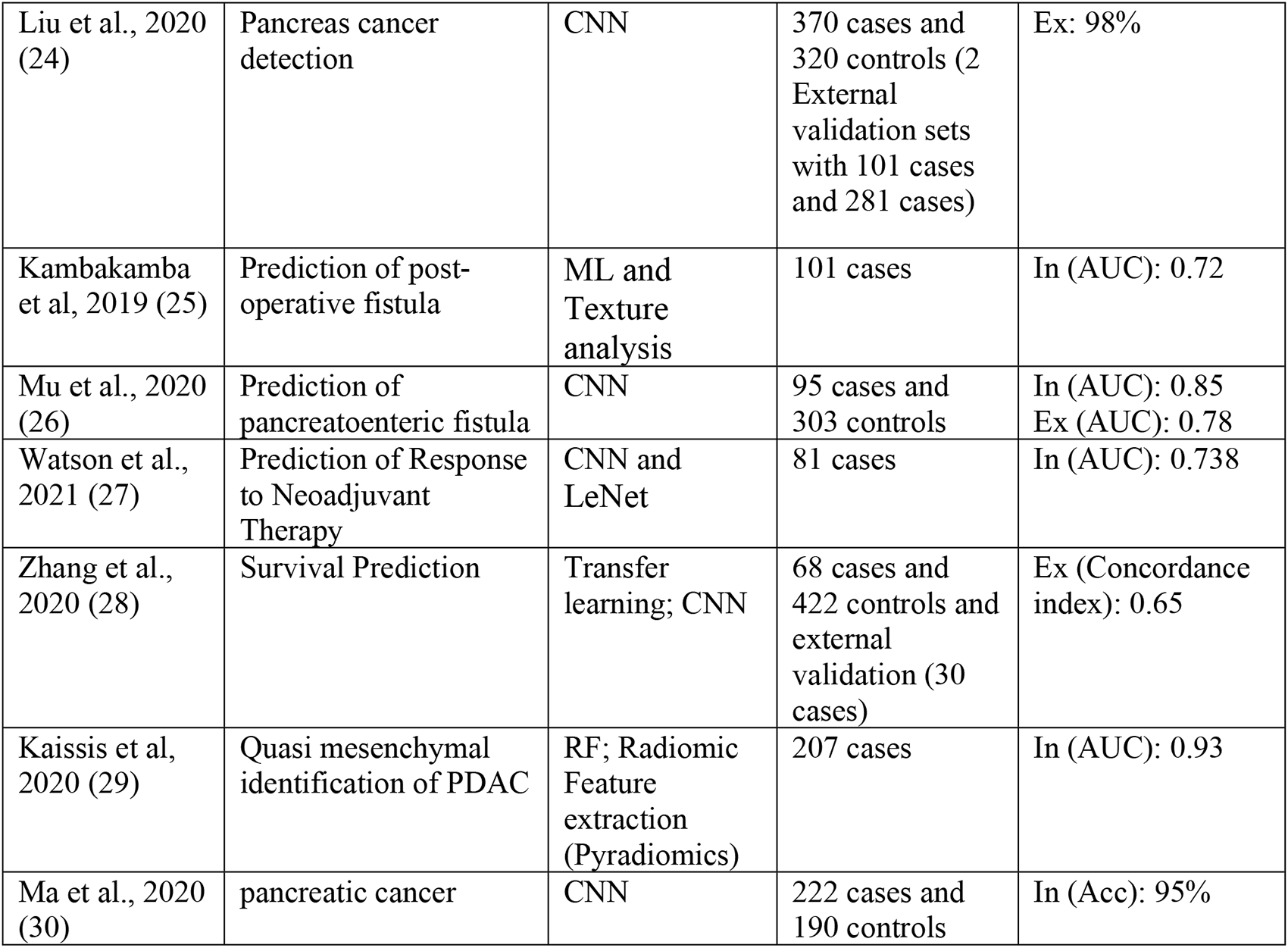
Literature of review for diagnostic and prognostic machine learning algorithms for pancreas CT scan imaging (22). Footnote: PDAC: Pancreatic ductal adenocarcinoma; RF: random forest; CNN: convolutional neural network; ML: Machine learning; Acc: Accuracy; AUC: Area under the receiver operating characteristic curve; Internal validation (In); External validation (Ex).

ML-guided tools’ design and desired outputs must be tailored toward implementation in healthcare systems (14). Thus, the current study describes the protocol for developing several computer-aided diagnoses (CAD) models to facilitate pancreatic cancer management using CT scan images. We sought to develop generalizable CAD systems to aid clinicians with a pancreatic cancer diagnosis (classification, segmentation, cancer subtype detection) and prognosis (cancer resectability, survival, staging) based on pancreas protocol CT scan images from 10 medical centers.

## Material and methods

### Ethical consideration

The Institutional Review Board of Research Institute for Gastroenterology and Liver Diseases (RIGLD), Shahid Beheshti University of Medical Sciences review board approved this ambispective study after consideration of data anonymization and security (code: IR.SBMU.RIGLD.REC.1401.043; link: https://ethics.research.ac.ir/EthicsProposalViewEn.php?id=323598). This protocol and future studies adhere to Helsinki Declaration of 1975, as revised in 2008, which provides ethical guidelines for medical research involving human subjects. We have taken necessary measures to protect the privacy and confidentiality of all participants and their personal data. Patients will be included prospectively from 1 December 2022 until March 2024 and retrospectively from 21 March 2015 to 23 October 2022. Informed consent will be collected through a phone call from the patient or their legal representative, providing a detailed description of the research aim and use. However, informed consent collection has been waived for patients collected retrospectively or in cases where access to the patient is not possible.

### Reporting guidelines and checklist

PanCanAID studies will be conducted adhering to the Standards for Reporting Diagnostic Accuracy (2015-STARD and STARD-AI) and Checklist for Artificial Intelligence in Medical Imaging (CLAIM) (31–33). The STARD checklist, flow diagram, and CLAIM is presented in **S1 Table**, **S1 Fig**, and **S2 Table**, respectively.

### Interdisciplinary Team Building

Starting in January 2021, a biweekly session was extended, with invitations sent to GI referral centers and healthcare providers. The study design involved a team of radiologists, gastroenterologists, surgeons, and computer science experts collaborating to develop the study.

Together, they discussed the data-gathering process, labeling techniques, and the desired machine learning tasks, which resulted in the current study design.

### Study design

This multicentric observational ambispective study will be conducted in ten medical centers in Tehran, Tabriz, and Guilan provinces of Iran: Taleghani Hospital in Tehran (T-H), Emam Khomeini Hospital in Tehran (EK-H), Firozgar Hospital in Tehran (F-H), Emam Hossein Hospital in Tehran (EH-H), Razi Hospital at Guilan province (R-H), Valiasr International Hospital at Tabriz province (V-H), Shariati Hospital in Tehran (S-H), Namazi Hospital at Shiraz Province (N-H), Shahid Faghihi Hosptial at Shiraz (SF-H), Behboud Specialized Clinic for Gastroenterology Diseases (B-C), and the Research Institute of Gastroenterology clinic (RIG-C). Patients will be included prospectively from 1 December 2022 until March 2024 and retrospectively from 21 March 2015 to 23 October 2022. An ethical review board waived gathering informed consent.

Patient demographic data will be collected from the electronic hospital information system (HIS). Image data will be obtained from the Picture Archiving and Communication Systems (PACS). EUS images will be obtained from a dedicated system (EndoPACS) at each local hospital system if available. CT scan and EUS images will be gathered in the “.dicom” and “.jpg” series. The medical team conducting the study will call all enrolled patients within two weeks of the enrollment to evaluate the survival and outcome of the cancer. **Fig 2** demonstrates the workflow and aims of PanCanAID.

**Fig 2.**
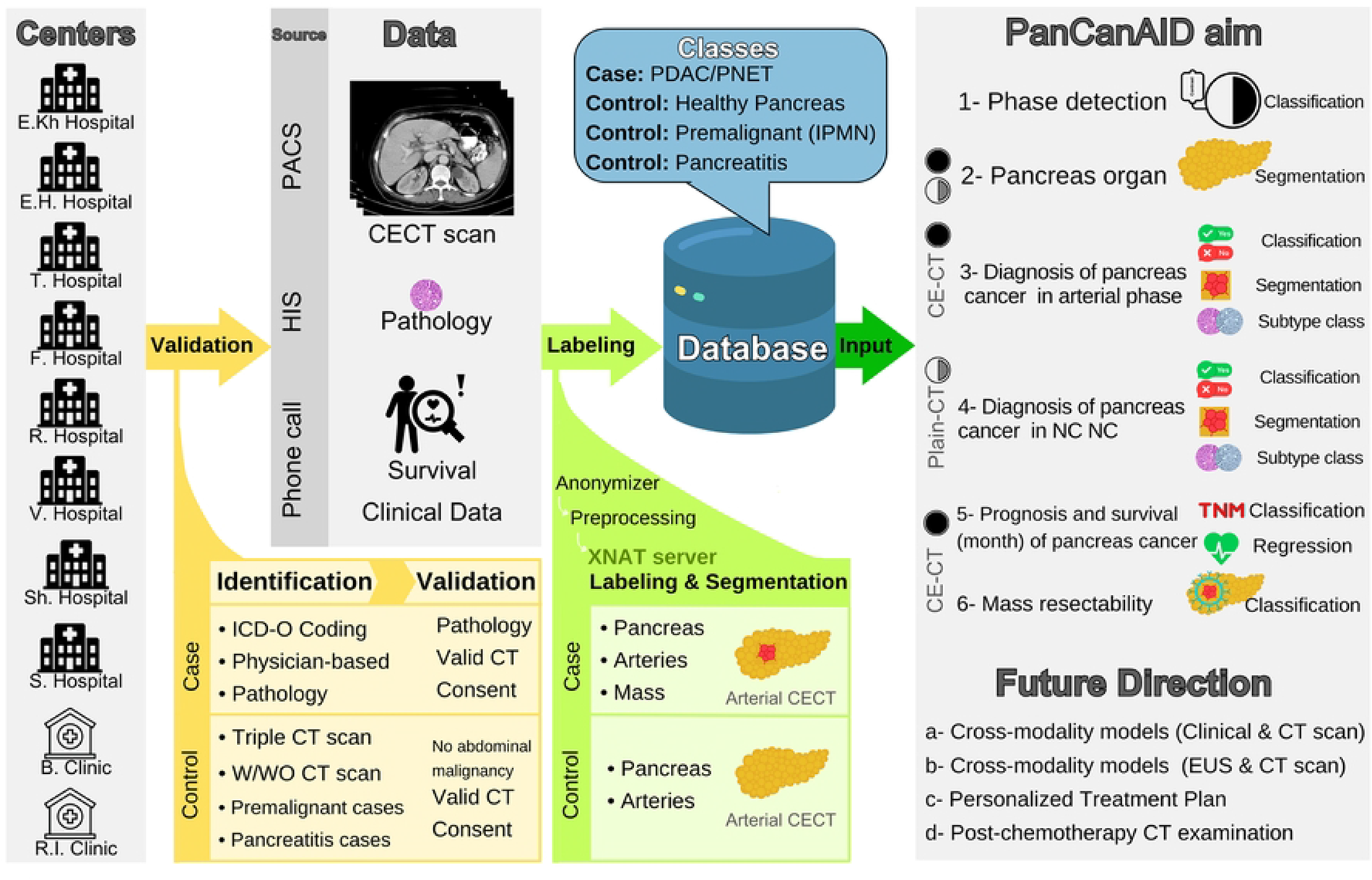
Workflow and aims of PanCanAID, a multicentric study to facilitate diagnosis and management of pancreas cancer. Footnote: The black circle represents a contrast-enhanced CT scan, and the half-black circle represents a non-contrast CT scan

### Patient eligibility, identification, and validation

Potential pancreatic cancer cases will be defined accord to using the following criteria: (1) international classification of diseases (ICD) code C25, (2) a histological diagnosis of pancreatic ductal adenocarcinoma (PDAC) OR pancreatic neuroendocrine tumor (PNET) or (3) a radiologic diagnosis of a pancreas mass OR pancreas tumor. Prospective enrollment of potential cases will occur at the time of referral to a radiology center or gastroenterology clinic.

Benign and premalignant lesions of the pancreas will be excluded from the case group. Patients without any valid CT scan before the initiation of treatment (chemotherapy or surgery) will be excluded. The treatment initiation will be obtained during a follow-up call or review of patient HIS records. Cases between 20 and 80 years old, with valid CT scan imaging and histologic confirmation of PDAC and PNET from pancreas specimens collected during surgery or FNA biopsy, will be included in the study. The suited CT scan for inclusion of cases and controls is triple phase CT scan or with and without contrast enhanced CT scan.

The control group will comprise three subgroups of patients aged 20 to 80 undergoing abdominopelvic CT scan and EUS examination but have no evidence of abdominopelvic malignancy or history of mass resection. The first subgroup will consist of patients without any pancreatic neoplasms. The second subgroup will consist of patients with premalignant lesions, primarily Intraductal papillary mucinous neoplasms (IPMNs), along with pancreatic intraepithelial neoplasia (PanIN) and mucinous cystic neoplasms (MCN) confirmed by pathology report (10). The third subgroup will consist of patients with acute or chronic pancreatitis confirmed by the radiologist. The selection of three subgroups aim to enrich the control group to represent the real-world challenge of diagnosing pancreatic cancer.

### Phone Interview for Survival and clinical data

Our prognostic model aims to predict the 6-month and 1-year survival of patients with pancreatic cancer. To achieve this, the medical team will conduct follow-up calls to both cases and controls, enrolled either retrospectively (over the past year) or prospectively, to collect survival time and relevant risk factors (10). The phone interview form used during these calls is attached in **S1 Appendix**. It includes information on the patient’s demographics, blood group, symptoms, diagnostic exams, treatments, smoking/alcohol consumption, diabetes/pancreatitis history, family history of cancer/pancreatitis, first symptom-diagnosis interval, and diagnosis-death interval. In addition to the phone interview data, two previously collected datasets from F-Hospital (with a 6-year follow-up) and Ekh-Hospital (with a 1-year follow-up) will be used, along with available imaging data retrieved from the PACS system.

### Sample size

Sample size estimation in machine learning projects in medical imaging requires an initial set of annotated data, which in our case, was unavailable (34). Moreover, a reliable method for estimating the sample size of biomedical ML research is unclear, especially with rapidly evolving modeling techniques (34). Figueroa et al. have proposed that between 80 and 560 annotated samples in each class are needed to achieve a root mean squared error lower than 0.01 (35). Similar studies using CT scan images achieved satisfactory results, with about 250 cases in each class of PDAC and non-PDAC (**Table 1**). The primary aim of PanCanAID is to collect data from 300 PDAC cases and 300 controls. We will examine the sample size using post hoc curve-fitting and the Figueroa method once the first 150 PDAC cases have been collected (75 cases and 75 controls) (35).

### Hospitals and imaging devises

Data will be collected from ten medical centers in Tehran. The center attributes, including the CT scanner model and weekly incidence of pancreatic cancer scans, are described in **Table 2**. Different imaging devices and technical variations may result in batch effect, so each patient’s imaging device and hospital sources will be recorded.

**Table 2.** Information of ten medical centers that will participate in patient enrollment. Footnote: PDAC: Pancreatic ductal adenocarcinoma; PNET: pancreatic neuroendocrine tumor.

| Hospital | GI referral center | Number of beds | CT scan Device | Estimated Workload of Pancreas cancer per week | Previously available data |
| --- | --- | --- | --- | --- | --- |
| Ekh-H | Yes | 2100 | 16 Detector Siemens SOMATOM Emotion | 15 | Yes (100 PNET Cases with CT scan) |
| EH-H | No | 500 | 16 Detector Siemens Somatom | 5 | No |
| T-H | Yes | 500 | 16 Decotor Siemenc Somatom | 10 | No |
| F-H | Yes | 554 | 16 Detector Siemens Somatom | 10 | Yes (200 PDAC patients with 2-year follow-up) |
| R-H | Yes | 500 | - | 7 | No |
| V-H | No | 1000 | - | 3 | No |
| Sh-H | Yes | 1000 | 2-MDCT Siemens Somatorm Volume Zoom, Siemens | 15 | No |
| S-H | Yes | 850 | 16-detector Somatom Emotion, Siemens | 7 | Yes (42 PDAC) |
| B-C | Yes | - | - | 3 | No |
| RIG-C | Yes | - | - | 5 | Yes (150 PDAC patients) |

### Online open datasets

We reviewed all previously published open-source pancreatic cancer CT imaging datasets, presented in **Table 3**. Previously segmented images from datasets such as WORD and AbdomenCT-1k will segment the pancreas on local images. This segmentation will be revised by the first investigator and confirmed by an experienced radiologist.

**Table 3.** Online open dataset for pancreas cancer CT scan imaging. Footnote: *Data used in this publication were generated by the National Cancer Institute Clinical Proteomic Tumor Analysis Consortium (CPTAC). Abbreviations: NCI: national cancer institute (of the United States of America)

| Name | Source | Data | License |
| --- | --- | --- | --- |
| AbdomenCT-1k (36) | GitHub (JunMa11/AbdomenCT-1k) | abdominal CT organ segmentation dataset with 1000+ CT scans by augmenting the existing single organ datasets | Apache-2.0 license |
| WORD (37) | GitHub (HiLab-git/WORD) | abdominal CT organ segmentation dataset | GNU General Public License v3.0 |
| Vindr (38) | Vindr.ai | Dataset of 1188 scans for phase recognition in abdominal contrast-enhanced CT | One can use the dataset without charge for non-commercial research purposes only |
| CPTAC-PDA (39) | The Cancer Imaging Archive (TCIA)* (40) | The NCI Clinical Proteomic Tumor Analysis Consortium collected CT scan images after pathological confirmation of 107 pancreas cancer cases | TCIA data usage policy |
| Pancreas-CT (41, 42) | The Cancer Imaging Archive (TCIA)* (40) | The National Institutes of Health Clinical Center performed 82 abdominal contrast-enhanced 3D CT scans in the portovenous phase | TCIA data usage policy |
| Pancreatic-CT-CBCT-SEG (43, 44) | The Cancer Imaging Archive (TCIA)* (40) | Breath-hold CT and cone-beam CT images with expert manual organ-at-risk segmentations from radiation treatments of 40 locally advanced pancreatic cancer patients | TCIA data usage policy |
| Our future dataset (PanCanAID) | The Cancer Imaging Archive or other imaging repositories | We plan to provide biphasic CT scans of 500 pancreas cancer cases with segmentation on arterial phase and patient outcome in 200 cases | GNU Affero General Public License v3.0 |

### Manual segmentation

We used a panel discussion and Chu et al.’s experience to decide on the segmentation strategy (17). Six radiologists will annotate and classify each axial plane of the abdominopelvic CT scan images in arterial phases of CECT using an approaches, an offline 3D slicer 5.0.3 program or an online XNAT server (45). The 3D slicer software will be used on a Windows-based local computer, and annotations will be made using lase pen (XP pen Deco 01 v2). “Brush” and “pen” tools will be used after setting the editable intensity Hounsfield range using the “threshold” tool. Using “threshold” will prevent selecting surrounding elements with different Hounsfield units. In the second approach, and for ease of access, an XNAT application on a Linux server with 200 Gb of storage and a two core 8Gb ram (46).

A second radiologist with expertise on abdominal imaging will review and confirm the annotations (not blinded to previous segmentations). In case of conflict, the data will be tagged as controversial, and conflicts will be resolved in a dedicated conflict resolution panel with two radiologists. Instruction for radiologists in the Persian language will be available before annotation, and five dedicated cases for educational purposes have been designed to ensure labeling and annotation uniformity.

### Active learning for segmentation

Providing ground truth annotations for medical images, especially in the case of pancreas images, is very time-consuming and requires limited expert resources. This is especially so for segmentation, where pixel-wise annotation is needed. Hence, we utilize active learning to interact with the annotator. Active learning is a technique in which a machine learning algorithm can improve its accuracy using less labeled training data by selecting the most informative data to learn from. Instead of being given a fixed set of labeled data, an active learner can ask an oracle to label additional instances that are most useful for improving its performance (47).

Active learning has been shown to be effective in radiology AI studies (48). We propose an automated system to carefully select the most representative data samples for annotation. We also consider the model’s uncertainty and approximate error probability on the new data sample for the selection. The selected samples are then given to the model for initial annotation. The annotated image is given to the radiologist to correct the annotation of the model and then added to the set of labeled datasets. After several steps, the newly labeled dataset is given to the model for retraining. We continue until the performance improvement stops or a pre-defined proportion of the dataset is labeled. The process is summarized in **Fig 3**.

**Fig 3.**
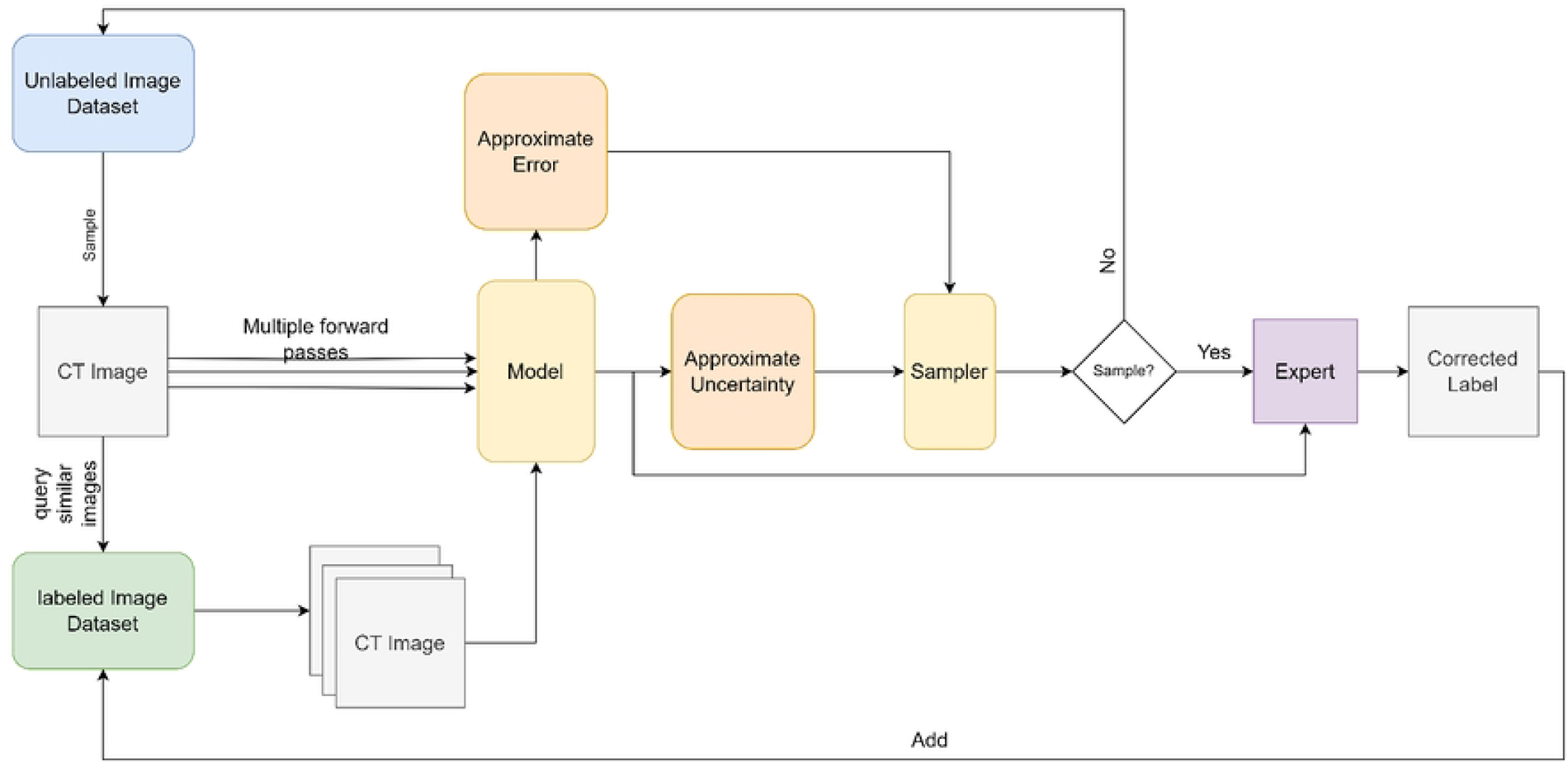
Workflow for manual segmentation using active learning approach.

### Mass characteristics

Radiologists will evaluate tumor characteristics, and this data will be used in the future phases of PanCanAID. These characteristics can be used to utilize automated reporting of pancreas mass in the future. **Table 4** shows pancreas cancer characteristics.

**Table 4.**
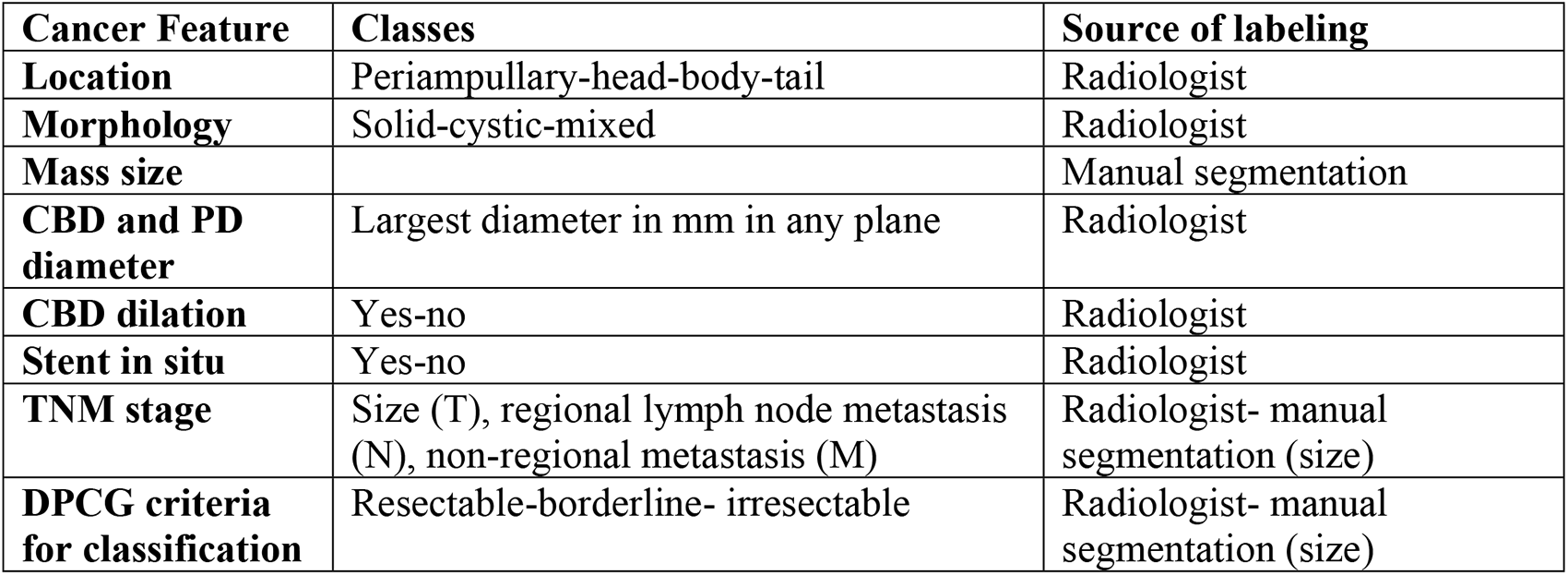
The routine report of mass characteristics and source of labeling in PanCanAID. Footnote: CBD: common bile duct; PD: pancreatic duct

| Cancer Feature | Classes | Source of labeling |
| --- | --- | --- |
| **Location** | Periampullary-head-body-tail | Radiologist |
| **Morphology** | Solid-cystic-mixed | Radiologist |
| **Mass size** |  | Manual segmentation |
| **CBD and PD diameter** | Largest diameter in mm in any plane | Radiologist |
| **CBD dilation** | Yes-no | Radiologist |
| **Stent in situ** | Yes-no | Radiologist |
| **TNM stage** | Size (T), regional lymph node metastasis (N), non-regional metastasis (M) | Radiologist- manual segmentation (size) |
| **DPCG criteria for classification** | Resectable-borderline- irresectable | Radiologist- manual segmentation (size) |

### Resectability definition and staging

The National Comprehensive Cancer Network (NCCN) and Dutch Pancreatic Cancer Group (DPCG) guidelines can help physicians in assessing the resectability of tumors (49, 50). This clinical decision is usually challenging for even expert radiologists. Both over-and undertreating can significantly impact a patient’s quality of life. We choose the DPCG criteria (**Table 5**) because of its simplicity and lower classification workload. An expert radiologist will classify CT scan images in the arterial phase, and this labeled data will be used to predict the resectability of mass. For TNM staging, a segmented pancreas mass will inform the tumor size (T-stage). A tumor with its longest diameter of less than 2 centimeters in an axial CT scan is defined as T1. Tumors with a diameter of 2-4 centimeters and those wider than 4 centimeters correspond to the T2 and T3 stages, respectively. The T4 stage consists of tumors involving vessels such as the celiac trunk, hepatic artery, and superior mesenteric artery. Radiologists will classify lymph node metastasis (N-stage) as a distant or regional invasion.

**Table 5.**
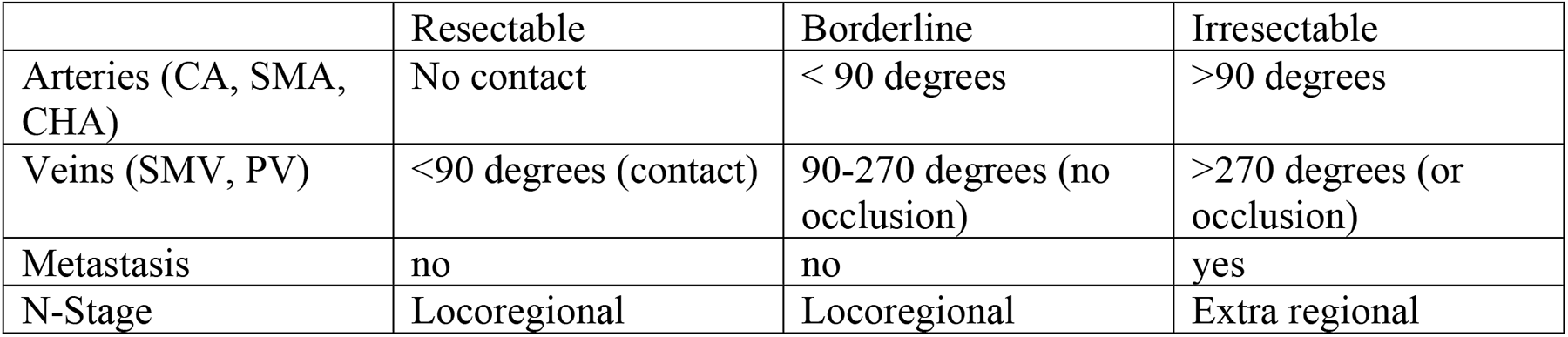
The Dutch Pancreatic Cancer Group (DPCG) criteria to assess the resectability of pancreatic cancer. Footnote: CA: celiac artery; SMA: superior mesenteric artery; CHA: common hepatic artery; SM: superior mesenteric vein; PV: portal vein.

|  | Resectable | Borderline | Irresectable |
| --- | --- | --- | --- |
| Arteries (CA, SMA, CHA) | No contact | < 90 degrees | >90 degrees |
| Veins (SMV, PV) | <90 degrees (contact) | 90-270 degrees (no occlusion) | >270 degrees (or occlusion) |
| Metastasis | no | no | yes |
| N-Stage | Locoregional | Locoregional | Extra regional |

### Data bank storage and computer processors

Data including “.dicom” files of CT scan images, “.jpg” image of EUS, “.csv” files with metadata (including patient characteristics, labels, hospital source, and notes as attached in Supplementary File 1), “.nrrd” files of 3D slicer with manual segmentations will be stored in three external hard drives with two terabyte storage. Processing will be handled using a pair of GTX 1080Ti GPUs. Performance and inference time will be evaluated on both GPU and CPU.

### CAD systems and tasks

As **Fig 2** depicts, different CAD models will be developed using a databank with specified aims, including:

1− Phase detection in abdominopelvic CT scan: classification (non-contrast phase, arterial phase, venous phase, portal phase, delay phase)
2− Pancreas organ segmentation (pancreas organ segmentation in CECT and non-contrast enhanced abdominopelvic CT scan)
3− Diagnosis of pancreas cancer in CECT scan: Classification (cancerous vs. non-cancerous), segmentation (pixel perfect pancreas organ and mass), and cancer subtype (PDAC and PNET) in CECT scan images (arterial phase)
4− Diagnosis of pancreas cancer in non-contrast CT scan: Classification (cancerous vs. non-cancerous), segmentation (pixel perfect pancreas organ and mass), and cancer subtype classification (PDAC and PNET) in non-contrast abdominopelvic CT scan images
5− Prognosis and survival of pancreas cancer in CECT: TNM stage classification including (T: size, N: lymph node metastasis, M: distant metastases), and survival (months) in contrast-enhanced CT scan images
6− DPCG resectability classification (resectable, borderline, and irresectable) in CECT scan images (arterial phase)

Future Direction: Multimodal approach for pancreas cancer ML tasks: Classification (cancerous vs. non-cancerous), cancer subtype (PDAC and PNET), resectability (resectable vs. irresectable), and survival (month) using demographics, EUS, and CT scan images

### Experiments and model development

Each input image is contrast-enhanced and denoised for all tasks. Normalization steps are conducted to mitigate the variability in data samples due to experimental conditions and measurement device configuration. For CT images, unnecessary slices are removed from the beginning and end of CT image sequences. We design and train a separate classification model that detects slices that have the pancreas visible, using the labels already gathered for our data. We consider slices with a minimum amount of detected pancreas area as positive and otherwise as negative. This filtering allows our models to be guided by useful cues and removes the computational cost of processing extra slices.

**Fig 4**. shows a brief overview of the model development workflow. The tasks are solved according to the following procedure:

**Fig 4.**
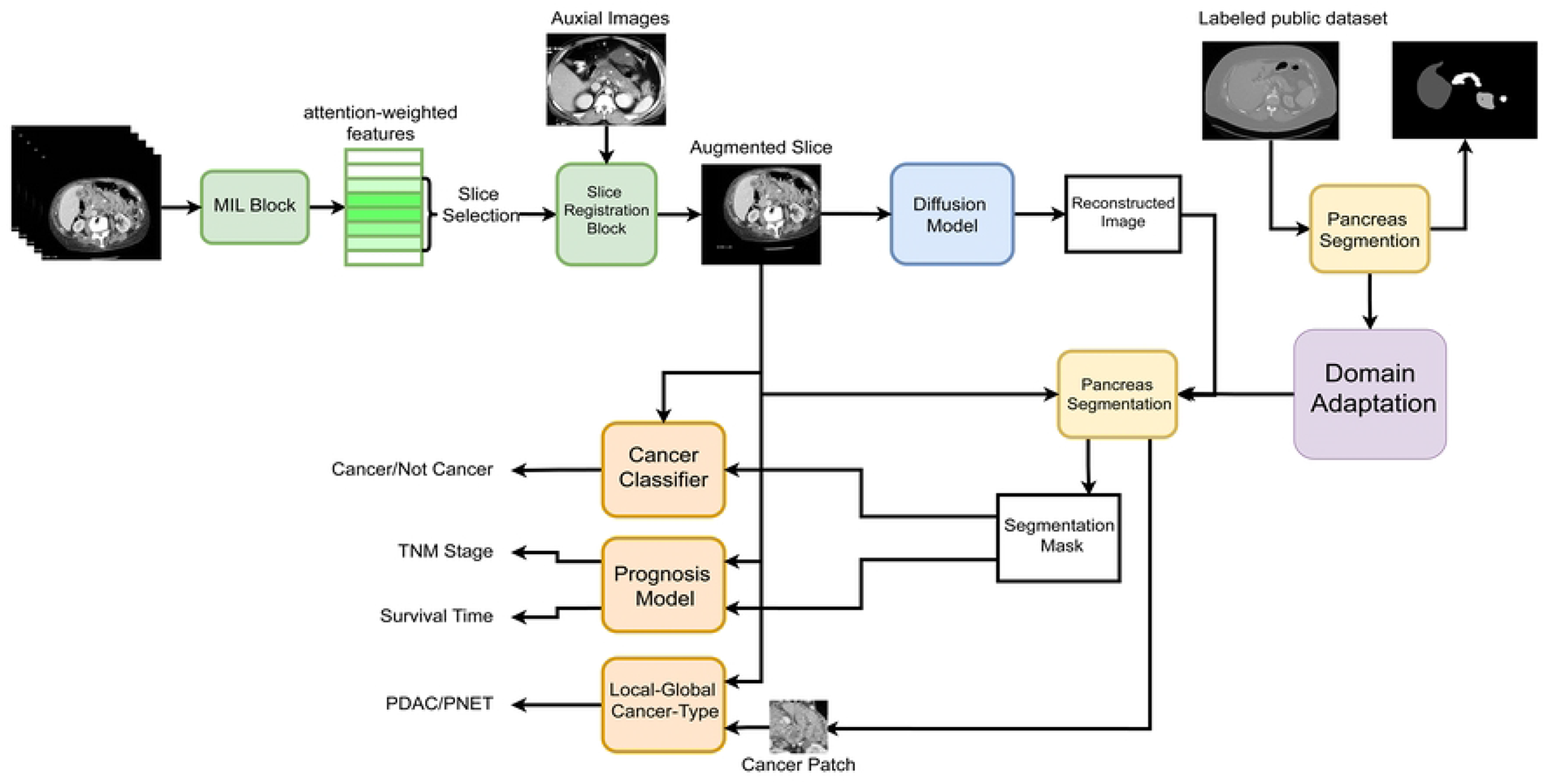
A brief overview of PanCanAID model development workflow.

1- A multi-instance learning approach is deployed to find substantial slices of data and make an initial detection of cancer. The features extracted in this step are also used for detecting and segmenting cancerous tissue in the following steps.

2- To extend our dataset, we conduct data augmentation. We use the variations between data samples to generate novel data. We train a spatial and an intensity registration network. Registration is a method to match images with the same structure but is distorted and hence not pixel-by-pixel matched (51). It finds a transformation that can map corresponding pixels to each other. We sample from the detected intensity and spatial maps learned from these models throughout the dataset and combine them to create new maps to generate new data.

3- A segmentation of the pancreas alongside any suspicious tissue mass is carried out in the CECT image. Due to the lack of labeled data and the cost of manual segmentation, we follow a domain adaptation approach. Domain adaptation has been extensively used in medical data and signals (52), especially in CT images (53)It is used when a model is supposed to be adapted to an unlabeled external domain with the help of the labeled internal dataset. We train a model on public datasets with segmentation labels and adapt the trained model to our dataset. We deploy different reconstruction methods to utilize the unlabeled data in our dataset. Reconstruction is mainly used to force latent features to contain as much information as possible to recreate the input data patterns (54). We gradually add labeled data and update the model according to an active learning framework to carry out the segmentation more accurately.

4- The cancer classification task uses segmentation from the original and reconstructed images. The delta image, the original image, and the features extracted in step 1 are processed through another Convolutional Network to detect cancer.

5- Upon cancer detection, we perform an additional classification task (PDAC or PNET) by processing a neighbor of the segmented mass in relation to the whole image. Local features are obtained by processing the area around the pancreas as well as the pancreas itself. Global features are computed based on the global attention of the whole image, considering the relationship between all regions in the image, providing a global representation of the structure of the abdominal CT image (55). We use local features around the segmented area and their relationship along with the global features of the entire image to model both the local information and the significance of this information relative to the global structure of the abdominal area. Local features

6- For the resectability classification task, we follow an approach similar to detecting the cancer type. We estimate the chance of successful tumor resection based on local-global feature extraction.

7- Segmentation estimation for plain non-contrast CT images is done by matching them to the label assigned to their corresponding CE-CT images. As we have segmentation maps for CE-CT images, we temporally align CT slices of patients with their CE-CT image to use the CE-CT labels as ground truth annotation for the plain CT images. The rest of the procedure is similar to the one for CE-CT images. Other tasks performed on the basic CT images are done similarly, as the true values of cancer labels, resectability, and prognosis results are the same for CE and plain CT images.

8- For prognosis, we model survival time as a conditioned normal variable, with mean and variance predicted by a 3D-CNN applied on the combination of the segmentation and original image. The mean head estimates the average survival time, and the (log) variance head estimates the uncertainty of the prediction. We infer mean and variance by maximizing the likelihood of the data.

### Batch effect removal

We designed a multi-level multi-site batch normalization (MMBN) architecture to remove the batch effect. We aim to remove batch effect at both the data- and feature-level.

### Data-level batch effect removal

To remove the effect of different measurement devices, we normalize the intensity and contrast of the CT images. We also apply affine normalization to remove geometrical biases caused by the experimental conditions.

### Feature-level batch effect removal

Only some of the variations in the data can be detected by low-level analysis of raw input images. We utilize our multi-site dataset to remove the feature-level batch effect that occurs in higher data abstractions. We deploy a Multi-Site Batch Normalization Layer (MSBNL) that consists of a batch normalization layer for each site in our dataset. The data sample is normalized according to the normalization parameters of its site. For a target site, we first estimate the mean and variance of the samples. Then, for each target-site sample, we pass it to each site-specific batch normalization layer and aggregate them using the weights defined as the KL divergence of the distribution of target site data and the corresponding source site data. To lessen the computational complexity, we assume a normal distribution for the data in each site. More concretely, assume site statistics (mean and variance) estimates from batch normalization parameters are (μ_*i*_, 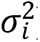) for each site-specific batch normalization layer and (μ*_t_*, 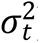) are the statistics of the target site. The weight of each layer for a sample from a target sample is calculated according to Equations 1 and 2:

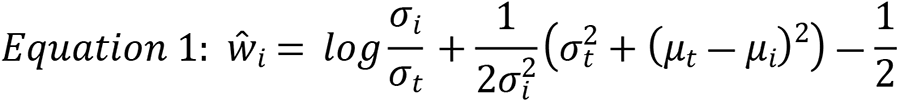

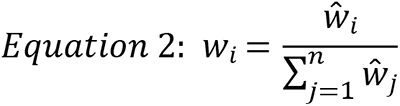

If we don’t have enough data from the target site, we also add the likelihood of the target data according to the distribution of each site using Equation 3:

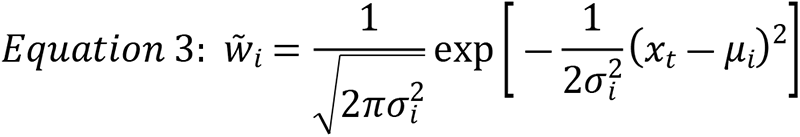

Where *x_t_*, is a feature of the sample from the target data, *u_i_* and *σ_t_* are expected value and variance of the feature for each domain according to the batch normalization layer, and *ŵ* = *ŵ* + *w̃* which means that we add computed *ŵ* parameters to *w̃* to account for a small amount of data in the target domain.

### Evaluation metrics and proposed model

The model performance will be tested in internal validation (test set) and external validation (from external hospital). For segmentation, we use IOU (intersection over union) to evaluate the proportion of detected mass. We also measure pixel-wise sensitivity and specificity to assess the model’s power to correctly find the true segmented areas and discard the unsegmented areas. F1 concludes these concepts as a single number. For classification (cancer and resectability), we measure the area under the receiver operating characteristic curve (AUROC), accuracy, sensitivity, specificity, and F1 by using the K-fold cross-validation technique (56). In addition, a calibration curve will be used to show how well the probabilistic predictions of a binary classifier are calibrated (57). We perform a simple statistical test to evaluate the prognosis’s predicted mean and variance values. For each ground truth prediction in the test dataset, we examine if it comes from the proposed normal distribution or not. The proportion of samples that pass the test is defined as a measure of the model’s performance.

### Explainable AI

The adaptation of AI models in the clinical setting has been constrained by their “black-box” nature, which makes it challenging for clinicians to comprehend and believe their predictions (58). Herby, we will embed Explainable AI (XAI) approach to increase its use case in the medical field. XAI is one of the branches of artificial intelligence concerned with building models that can offer clear and understandable justifications for their predictions and choices. By using architectures such as U-net, we can incorporate XAI techniques, such as Layer-wise Relevance Propagation (LRP) or Local Interpretable Model-agnostic Explanations (LIME), and we can provide visual explanations of the model’s predictions. Also, by segmenting the cancerous masses and pinpointing their location, medical professionals can better understand the reasons behind the model’s decisions and build trust in its predictions (58–60).

## Discussion

The development of CDSS and CAD tools requires interdisciplinary teamwork (17). PanCanAID is a multipurpose CDSS project addressing the current needs of pancreatic cancer care delivery across multiple phases of the disease. In developing the PanCanAID protocol, we addressed various aspects of team building, data collection and annotation, and model development. The process evolved over months of collaborative sessions with medical and computational experts. Many challenges were faced during protocol design, including the feasibility of data collection and annotation, complex multidisciplinary collaboration, optimization of data storage capacity, and the development of a state-of-the-art ML model. The protocol development process addressed these challenges and fostered effective cross-disciplinary collaboration toward a common goal.

CDSSs depend highly on a large, precisely-labeled dataset representing real-world data patterns (14). However, the truly adequate dataset size is unknown without conducting pilot studies (34). ML models of medical imaging may require even large datasets than models of tabular data due to the complexities of imaging data. The lack of publicly available cases on The Cancer Imaging Archive (TCIA) highlights the challenge of acquiring pancreatic cancer image data (**Table 3**). We constructed a multi-institutional team and designed it to collect sufficient cases for our models. Our initial aim is to gather 500 cases and 500 controls, but this number may be extended to reach the desired model performance: an AUC of 0.85 for diagnosis and 0.80 for cancer prognosis.

The data collection task for pancreatic cancer is more difficult for three reasons. First, a pancreatic cancer diagnosis is a relatively rare event (2). Second, the life span of patients is short, and many cases pass away within the first months, which makes patient identification even harder (1, 61). The third and most important reason is the winding journey of pancreatic cancer patients during diagnosis and management. Multiple medical centers and specialties manage pancreatic cancer, and the data is stored in various sources (**Fig 5**). The collection of this data needs rigorous amounts of time and a substantial amount of effort (17). We used an ambispective design to collect more accessible cases retrospectively and precise data prospectively. However, finding pathologically confirmed patients with survival outcomes can be unattainable in many cases. We aim to collect the survival of patients, which is needed for some tasks, by calling patients diagnosed in the last year or included prospectively.

**Fig 5.**
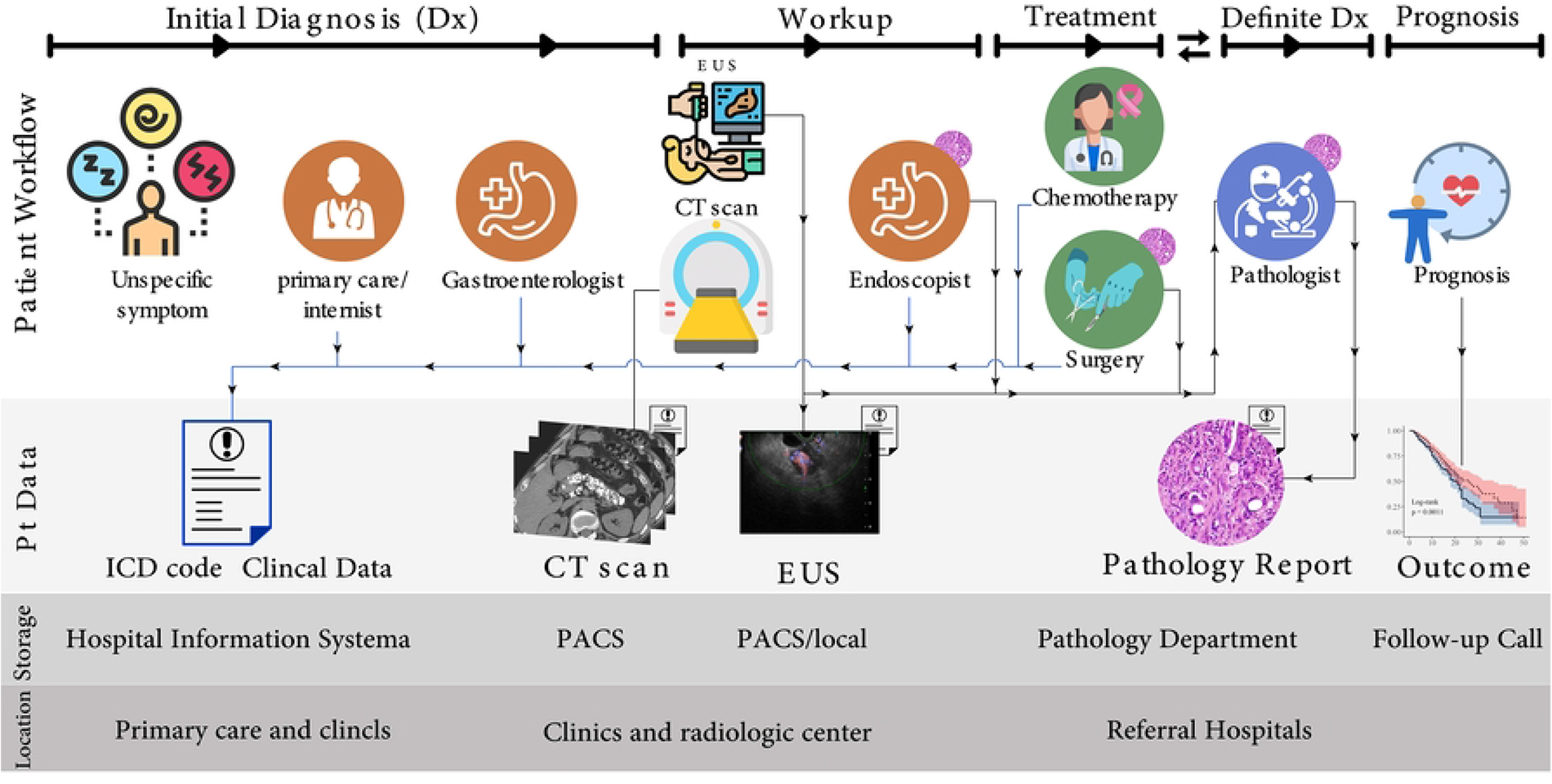
Dissemination, storage, patient data sources, and the journey of patients with pancreas cancer.

The next issue is the time and effort needed to label the data. In our case, tumor characteristics, pancreas segmentation, tumor segmentation, and resectability assessment were warranted. We assembled a team of junior radiologists with six years of experience who will handle segmentation. An active learning model will also be developed using publicly available data for pancreas organ segmentation and less than 100 local cases. Two senior radiologists who are experts in the field will validate and confirm the labeled data. The interobserver variability will be reported to show the use case of CADs.

In addition, we sought to collect other data, such as EUS images and clinical data (symptoms, past medical history, and demographics). We will use the collected data to develop a cross-modality platform, which is the future direction of PanCanAID. This data bank can overcome the current bottleneck in the model development of pancreatic cancers.

Our project involves a comprehensive benchmarking of previous models and the development of new algorithms. However, several challenges make our task more complex than a typical classification/segmentation task. Firstly, labeling all CT scans, particularly those with narrow imaging cuts, is impractical due to the high cost of segmentation labeling. To overcome this, we have adopted an active learning approach where radiologists interact with our team to validate and correct segmentation labels. This results in higher quality annotations and improved sample efficiency. Secondly, the complex structure of abdominal images, especially the irregular shape of the pancreas, can benefit from large-scale external data with extensive labeling. We aim to deploy a domain adaptation framework to transfer the knowledge learned from the large-scale external data into our dataset. Thirdly, we have designed a batch-effect removal protocol to eliminate batch effects in our multi-source data, which we will extend using other domain generalization techniques. Finally, we aim to build our model with unlabeled data, using semi-supervised learning as a key component of our framework.

Although we tried to overcome several challenges during the protocol design process, several unknown factors could still affect our future work. The presence of all three needed data modalities (CT scans, pathology reports, and survival data) may be unachievable in many pancreatic cancer cases. In addition, the quality of CT scan images, especially in the arterial phase, may be insufficient. Developing a generalizable model will need rigorous effort for batch effect removal. In addition, the segmentation and labeling of data require a vast amount of time from radiologists, which may be exhausting. We hope to overcome upcoming challenges through interdisciplinary teamwork.

## Conclusion

PanCanAID is a large-scale AI project developing CADs and CDSSs using pancreatic cancer CT scan images. In hopes of improving pancreatic cancer prognosis, it will tackle the current bottleneck of model development and data shortage. We plan to collect good quality, sufficient amounts, and precisely labeled data banks by creating a team of experts from various institutions. Besides, in our model development, we utilize and expand different concepts according to the challenges we have in our task, including active learning, semi-supervised learning, and domain adaptation and generalization. Experts in medical and computational fields were involved in protocol development, striving to describe the problem from all aspects. The protocol design lasted for months, but it fostered the replicability of the method and cross-disciplinary teamwork.

## Data Availability

The model will be published for non-commercial future use of researchers under certain license. The data will be published and stored in a renowned repository for future non-commercial use of researcher.

## Acknowledgment

The authors want to thank the National Elite Foundation, Research Institute of Gastroenterology and Liver Diseases (Shahid Beheshti University of Medical Sciences), Data Science and Machine Learning (DML) Lab (Sharif University of Technology), Advanced Diagnostic and Interventional Radiology Research Center (Tehran University of Medical Sciences), Gastroenterology and Liver Diseases Research Center (Iran University of Medical Sciences), Gastrointestinal and Liver Diseases Research Center (Guilan University of Medical Sciences), and Medical Imaging Research Center (Shiraz University of Medical Sciences) for providing resources and support.

## Supporting Information

**S1 Table.** The Standards for Reporting Diagnostic Accuracy (STARD 2015)

**S2 Table.** Checklist for Artificial Intelligence in Medical Imaging (CLAIM)

**S1 Figure.** STARD 2015 patient flow diagram for PanCanAID

**S1 Appendix.** The phone interview form for collecting survival and clinical data

